# Processes of Spatial Stratification: A Scoping Review Protocol

**DOI:** 10.1101/2025.02.24.25321457

**Authors:** Dominika Dmitrzak

## Abstract

**Objective:** The objective of this scoping review is to understand what is known and theorised about the processes of spatial stratification.

**Introduction:** Regional inequalities, commonly termed the North-South divide, have been an feature in England for years(1). Whilst there is research on the different areas affected, these include, but not limited to, income(2), education(3), and health(4), there is a lack of research on the processes that lead to the differentiation of regions, in other words, on spatial stratification. The aim of this scoping review is, therefore, to identify and synthesise what is known about the processes and factors involved in spatial stratification, bringing together research from separate fields such as economics, sociology, and geography.

**Eligibility Criteria:** In line with standard scoping review approaches, Population, Concept, Context (PCC)(5) will be used as a guide for eligibility. Population: anyone; Concept: spatial or socio-spatial stratification; Context: any country, any level. Articles and papers written in English, published after 1970, and meeting the inclusion criteria outlines will be included in the review.

**Methods:** Searches will be conducted in Scopus, Web of Science, JSTOR, Embase, and ProQuest Social Science Premium Collection. The time frame for the searches will be set from 1970 onwards. All titles and abstracts will be screened against the inclusion criteria in Rayyan. Reference lists of all eligible papers will be searched for further relevant literature. Full-text versions of eligible literature will be reviewed and data relating to theories, concepts, and where relevant, findings, will be extracted. Papers meeting the inclusion criteria will be tabulated, grouped thematically, and written up to form a narrative describing what is currently known and theorised about the processes that lead to spatial stratification.

## Introduction

Regional inequalities, commonly thought of as the North-South divide, have been documented in England for years(1). Although they are far from a new phenomenon, disparities between regions have widened in the 21^st^ century and the UK now has the largest regional inequalities of any comparable country (1,6).

These regional inequalities can be observed across multiple measures and extend to many aspects of daily life. A popular focus amongst economists is the difference in economic productivity, which is above UK average in London and the South East and below average in Northern regions (2). A similar disparity can be seen with household income(7). Aside from impacts on the country’s economy, this is important as over time, a region’s economic productivity is a key factor in it’s standards of living(7).

However, regional inequalities do not stop at income and economic productivity. The life expectancy gap in 2020-2022 between the South East, the region with the highest life expectancy, and the North East, the region with the lowest life expectancy, was 3 years. Comparing the most deprived areas in England with the least deprived areas, the gap in life expectancy is over 10 years for men and over 7 years for women (4). The North experiences above average infant mortality rates and reports higher rates of bad health and disability than average. Northern regions experience higher rates of economic inactivity due to ill health or disability. In fact, the top five local authorities with the highest levels of economic inactivity due to long-term illness or disability are all in the North(8).

Whilst there is research on the different areas affected by regional inequalities, as mentioned these include, but are not limited to, income(7), education(3) and health(4), there is a lack of research on the processes that lead to the differentiation of regions, in other words, on spatial stratification. The aims of this review are to identity and synthesise literature on this topic.

Rather than containing empirical evidence, part of the literature on spatial stratification is conceptual, therefore a scoping review has been deemed more appropriate than a systematic review. Furthermore, the aim of the review is not to find answers to a specified question, in which case a systematic review would be appropriate, but to identify the scope of the literature and the different concepts, theories and frameworks used to explain the processes of spatial stratification across multiple disciplines, for which a scoping review is a more appropriate choice(9).

## Research Question

What is known and theorised about the processes of spatial stratification?

### Study Design

Scoping review methodology will be used to identify and draw together literature on the processes and theories behind spatial stratification from across disciplines. The review will be conducted following an established framework (5,10,11) and will adhere to the Preferred Reporting of Systematic Reviews extension for Scoping Reviews (PRISMA-ScR)(12). The PRISMA checklist has been used to develop this protocol (see Appendix 1).

### Search Strategy

The PRISMA-ScR guidelines(12) will be used during the design, conduct, and reporting of the review. Using identified key words, a search of multidisciplinary research databases (Web of Science, Scopus, JSTOR, Embase, and ProQuest Social Science Premium Collection) will be conducted. References lists of eligible material will also be searched. To ensure a wide scope, the search will include literature published from 1970 onwards. Literature published before this time will not accurately reflect the political and economic climate and will be excluded from this review. Three articles meeting the inclusion criteria were identified and used to ensure sensitivity of searches across databases (Table 1).

**Table 1:** Sensitivity Papers.

| **Table 1: Sensitivity Papers** |  |
| --- | --- |
| **Reference** | **Retrieved in pilot searches** |
| Logan (1978) Growth, Politics, and the Stratification of Places (13) | Yes |
| Wu & Li (2005) Sociospatial Differentiation: Processes and Spaces in Subdistricts of Shanghai (14) | Yes |
| Kühn (2015) Peripheralization: Theoretical Concepts Explaining Socio-Spatial Inequalities (15) | Yes |

### Eligibility Criteria

In line with standard scoping review approaches, Population, Concept, Context (PCC)(5) will be used as a guide for eligibility (see Table 2). The time frame of the search will be from 1970 – 2024. Although this scoping review is part of a project focusing on England, the review will not be limited to England, alone. Country specific context is important to acknowledge, however, this limit would risk missing key insights. Moreover, preliminary searches show that imposing this limit would narrow down the pool of literature to a degree incompatible with performing a meaningful review. To understand the full scope of literature on the topic, journal articles and conference papers will all be included in the review. For literature which cannot be accessed via institutional holdings, the source authors will be contacted to assist with procuring access. Literature on the processes, causes, and factors involved in spatial and/or socio-spatial stratification will be included in the review. Due to time and cost involved in translating material, only material in English will be included in the review.

**Table 2:** PCC guide.

| Table 2: PCC guide |  |
| --- | --- |
| Population | All ages, ethnicities, and nationalities |
| Concept | Spatial or socio-spatial stratification |
| Context | Any country, any level |

### Screening

Titles and abstracts will be screened in Rayyan (https://www.rayyan.ai/) and relevant papers will be retrieved and assessed for inclusion. Following the PRISMA guidelines, a flowchart for the inclusion process will be produced. Literature will be selected based on the inclusion criteria outlines in Table 3.

**Table 3:** Inclusion & Exclusion Criteria.

| Table 3: Inclusion & Exclusion Criteria |  |
| --- | --- |
| Inclusion Criteria | Exclusion Criteria |
| Articles or paper is written in English. | Articles or paper is not written in English. |
| Article or paper has been published from 1970 onwards. | Article or paper was published before 1970. |
| Article or paper discusses processes, causes, and/or factors involved in spatial and/or socio-spatial stratification. | Article or paper does not discuss processes, causes, and/or factors involved in spatial and/or socio-spatial stratification. |

### Data Extraction

Data relating to theories, concepts, and where relevant, findings, will be extracted from full-text versions of included literature (see Appendix 3). The aim of a scoping review is not to find answers to a specified question, but rather to map out the scope of available literature on the topic(5). As such, an assessment of the quality of literature is not typically undertaken and will also not be conducted in this review.

### Synthesis

Papers meeting the inclusion criteria will be tabulated, grouped thematically, and written up to form a narrative describing what is currently known and theorised about the processes and factors that contribute to spatial stratification.

## Data Availability

N/A

## Author Contact

Dominika Dmitrzak, The School of Geography, Politics and Sociology, Newcastle University, Newcastle, UK.

---

**Appendix 1:** PRISMA-ScR checklist.

| Section and topic | Item No | Checklist item | (Page No.#) |
| --- | --- | --- | --- |
| **ADMINISTRATIVE INFORMATION** |  |  |  |
| Title: |  |  |  |
| Identification | 1a | Identify the report as a protocol of a systematic review | 1 |
| Update | 1b | If the protocol is for an update of a previous systematic review, identify as such | N/A |
| Registration | 2 | If registered, provide the name of the registry (such as PROSPERO) and registration number | N/A |
| Authors: |  |  |  |
| Contact | 3a | Provide name, institutional affiliation, e-mail address of all protocol authors; provide physical mailing address of corresponding author | 4 |
| Contributions | 3b | Describe contributions of protocol authors and identify the guarantor of the review | 4 |
| Amendments | 4 | If the protocol represents an amendment of a previously completed or published protocol, identify as such and list changes; otherwise, state plan for documenting important protocol amendments | N/A |
| Support: |  |  |  |
| Sources | 5a | Indicate sources of financial or other support for the review | 4 |
| Sponsor | 5b | Provide name for the review funder and/or sponsor | 4 |
| Role of sponsor or funder | 5c | Describe roles of funder(s), sponsor(s), and/or institution(s), if any, in developing the protocol | 4 |
| **INTRODUCTION** |  |  |  |
| Rationale | 6 | Describe the rationale for the review in the context of what is already known | 1-2 |
| Objectives | 7 | Provide an explicit statement of the question(s) the review will address with reference to participants, interventions, comparators, and outcomes (PICO) | 2 |
| **METHODS** |  |  |  |
| Eligibility criteria | 8 | Specify the study characteristics (such as PICO, study design, setting, time frame) and report characteristics (such as years considered, language, publication status) to be used as criteria for eligibility for the review | 3 |
| Information sources | 9 | Describe all intended information sources (such as electronic databases, contact with study authors, trial registers or other grey literature sources) with planned dates of coverage | 2 |
| Search strategy | 10 | Present draft of search strategy to be used for at least one electronic database, including planned limits, such that it could be repeated | Appendix 2 |
| Study records: |  |  |  |
| Data management | 11a | Describe the mechanism(s) that will be used to manage records and data throughout the review | 3 |
| Selection process | 11b | State the process that will be used for selecting studies (such as two independent reviewers) through each phase of the review (that is, screening, eligibility and inclusion in meta-analysis) | 3 |
| Data collection process | 11c | Describe planned method of extracting data from reports (such as piloting forms, done independently, in duplicate), any processes for obtaining and confirming data from investigators | 3 and Appendix 3 |
| Data items | 12 | List and define all variables for which data will be sought (such as PICO items, funding sources), any pre-planned data assumptions and simplifications |  |
| Outcomes and prioritization | 13 | List and define all outcomes for which data will be sought, including prioritization of main and additional outcomes, with rationale | Appendix 3 |
| Risk of bias in individual studies | 14 | Describe anticipated methods for assessing risk of bias of individual studies, including whether this will be done at the outcome or study level, or both; state how this information will be used in data synthesis | N/A |
| Data synthesis | 15a | Describe criteria under which study data will be quantitatively synthesised | N/A |
| | 15b | If data are appropriate for quantitative synthesis, describe planned summary measures, methods of handling data and methods of combining data from studies, including any planned exploration of consistency (such as $I^2$, Kendall's $\tau$) | N/A |
|  | 15c | Describe any proposed additional analyses (such as sensitivity or subgroup analyses, meta-regression) | N/A |
|  | 15d | If quantitative synthesis is not appropriate, describe the type of summary planned | 4 |
| Meta-bias(es) | 16 | Specify any planned assessment of meta-bias(es) (such as publication bias across studies, selective reporting within studies) | N/A |
| Confidence in cumulative evidence | 17 | Describe how the strength of the body of evidence will be assessed (such as GRADE) | N/A |
| Selection of sources of evidence† | 9 | State the process for selecting sources of evidence (i.e., screening and eligibility) included in the scoping review. | 3 |
| Data charting process‡ | 10 | Describe the methods of charting data from the included sources of evidence (e.g., calibrated forms or forms that have been tested by the team before their use, and whether data charting was done independently or in duplicate) and any processes for obtaining and confirming data from investigators. | 3 and Appendix 3 |
| Data items | 11 | List and define all variables for which data were sought and any assumptions and simplifications made. | Click here to enter text. |
| Critical appraisal of individual sources of evidence§ | 12 | If done, provide a rationale for conducting a critical appraisal of included sources of evidence; describe the methods used and how this information was used in any data synthesis (if appropriate). | N/A |
| Synthesis of results | 13 | Describe the methods of handling and summarizing the data that were charted. | 4 |

**Appendix 2:** Search strategy in Scopus (the same strategy will be followed in Web of Science and JSTOR)

| Search Terms | Results from<br>08/07/2024 |
| --- | --- |
| "place stratification" OR "stratification of place" OR "stratification of space" OR "neighbourhood stratification" OR "socio-spatial stratification" OR "socio-spatial differentiation" OR "sociospatial differentiation" OR "socio-spatial differentiation" AND process OR "neighbourhood inequalit*" OR "internal migration" AND "spatial inequalit*" OR "internal migration" AND "social class" OR "socio spatial division" OR "socio-spatial division" "residential differentiation" OR "residential differentiation" AND migration OR "residential differentiation" AND (space OR place OR spatial) OR "internal migration" AND (selective OR selection OR selectivity) "social cleansing" OR "socio-spatial polarisation" OR "socio-spatial polarization" OR "residential segregation" AND process OR "sociospatial structure" OR "socio-spatial structure" OR peripheralization OR peripheralization OR "socio spatial development" AND uneven OR "regional inequalit*" AND "uneven development" OR gentrification AND "spatial inequalit*" OR "spatial sorting" OR "social mobility" AND "spatial mobility" OR "socio-spatial differentiation" AND mobility OR "socio-spatial stratification" AND mobility OR "social mobility" AND "spatial stratification" | 5718 |
| Limit : 1970 – current | 5704 |

**Appendix 3:**
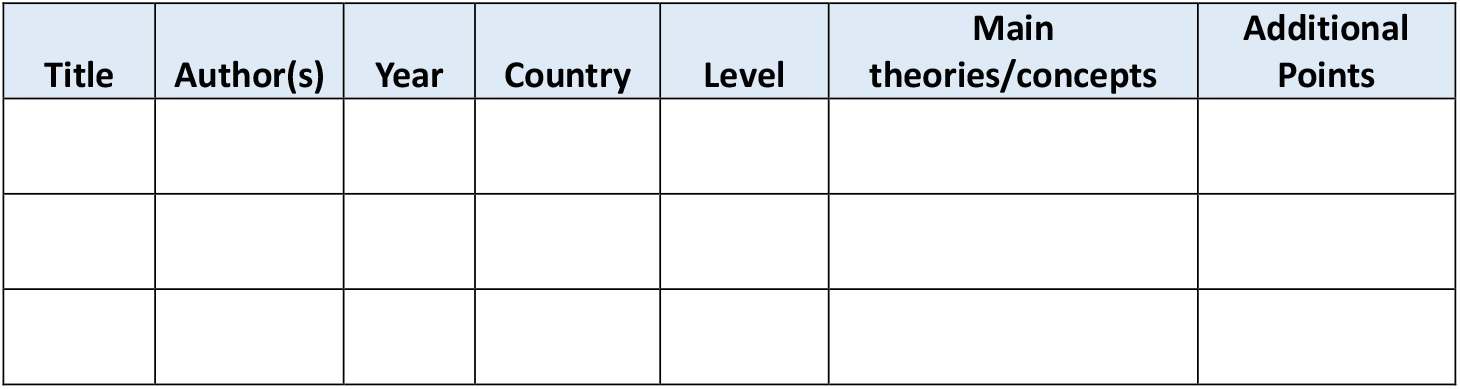
Data extraction template.

